# Chronic suppurative otitis media: understanding its prevalence, bacterial spectrum, and antimicrobial resistance patterns among patients presenting with middle ear infections at KCMC in 2022, Tanzania: A hospital-based crosssectional study

**DOI:** 10.1101/2025.06.11.25329461

**Authors:** Simon Manyata, Dhahiri Mnzava, Pius Tarimo, Hosiana J. Mo, John E. Nyigu, David J. Opundo, Ezekiel B. Gamuya, Adam Mwakyoma, Debora C. Kajeguka

## Abstract

**Background:** The global incidence of chronic suppurative otitis media is as high as 4.76% i.e. 31million cases with 22.6% of these cases occurring in children under five years. With a prevalence of 30.82 per ten-thousand, each year 21 thousand people die due to complications of chronic suppurative otitis media.

**Objective:** the objective of this study was to determine the prevalence of chronic suppurative otitis media, its bacteriological profile, and antimicrobial susceptibility pattern among patients presenting with middle ear infections at KCMC, 2022.

**Methodology:** This hospital-based cross-sectional study was conducted at KCMC, ENT department from 15-03-2022 to 30-05-2022 and enrolled 201 participants by convenient sampling. From those confirmed with CSOM, a discharge swab was collected for microbiological investigations. Antimicrobial susceptibility was performed using the Modified Kirby-Bauer disk diffusion method. Data was analyzed using SPSS.

**Results:** Of 201 participants, 23 (11.4%) were confirmed to have CSOM. Prevalence of CSOM was higher at 22.4% (13/58) in children under five years of age. The proportion of CSOM infection was higher among males 12.9% (11/85) compared to females 10.3% (12/116). CSOM prevalence was high among rural participants 15.7% (17/108) compared to urban 6.4% (6/93). Out of 23 CSOM discharge swabs, 82.6% (19/23) were culture-positive. *S. aureus* was the most common isolate 28.57% (6/23). Most isolates were Multi-drug resistant with the highest drug resistance found against penicillin antibiotics (60%-70%), lower to medium resistance (11-50%) against Cefotaxime, Meropenem, Ciprofloxacin, Chloramphenicol, Ceftazidime, Erythromycin, and Cefoxitin. At the same time, there was no resistance to Amikacin.

**Conclusion:** The high prevalence of CSOM and a high proportion of MDR bacteria among CSOM patients is a threat to patient management. Continuing empirical treatment could escalate the problem. We highly recommend microbiological analysis of middle ear discharge to study bacterial profile and drug susceptibility for the provision of proper and effective antibiotic treatment to CSOM patients.

## INTRODUCTION

Chronic suppurative otitis media (CSOM) is a chronic middle ear infection characterized by a persistent discharge through a tympanic perforation for two weeks and above (1–5). CSOM in most cases begins in childhood as spontaneous tympanic perforation due to untreated acute infection of the middle ear, this is known as acute otitis media (AOM) (6,7). Complications of CSOM are of great concern if not well managed (7,8). As far as we understand, infection of the middle ear is among the preventable causes of hearing loss and impairment in people of all age groups, more observed in children population (9–12). The global incidence of CSOM is as high as 4.76%, with more cases 22.6% occurring in children under five years (13). In Africa, the overall prevalence is 6% (14). Tanzania is estimated to be among the countries with the highest proportion of CSOM with an overall prevalence of 14% (7). Poor health systems and Laboratory investigations have made CSOM endemic in tropical and sub-Saharan Africa (15).

The common causatives of CSOM are bacterial pathogens followed by viruses and fungi (16,17). The bacterial agents of chronic suppurative otitis media are *Pseudomonas aeruginosa, Staphylococcus aureus, Klebsiella pneumoniae, Escherichia coli, Proteus spp, Streptococcus pneumoniae, Hemophilus influenza, Morexella carrhalis* and *Coagulase negative Staphylococci* (18–20). However, the proportions and bacterial profile of CSOM causatives vary across various geographical locations, social groups, and levels of development. There is a great knowledge gap in the microbiology of CSOM especially in developing countries due to a lack of proper Laboratory investigation (21). Repeated empirical prescription of antimicrobials and overuse of antibiotics without prior laboratory identification of bacterial species and performance of antimicrobial susceptibility testing are among the causes of gene mutation and selection pressure in the community- and hospital-acquired pathogens which makes them drug/multidrug-resistant (22,23).

Antimicrobial resistance (AMR) is a rapidly growing global burden and imposes a barrier to the treatment of various bacterial diseases (24). AMR is attributed to various factors including poor knowledge, awareness of the rational use of antimicrobials, and self-treatment (21,25,26). In developing countries, a cocktail of antimicrobial ear drops is prescribed without any prior microbiological analysis of ear discharge (27). In Tanzania, CSOM patients are prescribed ciprofloxacin and ofloxacin ear drop antibiotics (28). Due to poor antimicrobial stewardship, CSOM pathogenic bacteria are increasingly becoming resistant to basic and accessible antimicrobial agents (10,29). Studies report the resistance of CSOM bacterial isolates to widely prescribed ciprofloxacin (24,30). Resistance to topical quinolones is an alarming threat to the public including Tanzania where it is among the highly prescribed antibiotic for CSOM patients (7,28). There is a strong association between prolonged antibiotic use and the selection of AMR (31), this confirms that microbiological samples need to be tested before antibiotic prescription.

## Materials and Methods

### Study design and settings

This was a hospital-based cross-sectional study conducted from 15-03-2022 to 30-05-2022. The study was conducted at KCMC, the only hospital with a fully established Ear Nose and Throat (ENT) department in north Tanzania. KCMC receives referred patients, the majority of them are from northern zone regions i.e., Arusha, Manyara, Tanga, and Kilimanjaro, and bordering regions of countries like Kenya. The hospital has a capacity of more than 600 beds and receives hundreds of patients per day. KCMC/ENT clinic attends more than 10,000 patients with Otitis media annually. The hospital’s ENT department was an ideal location for this study due to its significant patient volume, experienced medical personnel, and comprehensive services provided.

### Ethical consideration

Ethical approval for the study was obtained from the Kilimanjaro Christian Medical University College Research Ethics Review Committee (CRERC) with certificate No. PG 12/2022 (see appendix xiv). Permission for data collection and laboratory analysis was sought from the Head of the ENT Department KCMC and the Head of the Head of the Health Laboratory Sciences Department KCMC University. Eligible participants were provided with a written or audio (a researcher read) form of informed consent explaining the aim of the study, the impact of the study on society, and assurance of confidentiality. For participants below 18 years of age, their parents or guardians read or listened to the informed consent, explained to their children, and if they agreed to participate a parent/guardian signed for them. It was also assured to them that their refusal to participate in the study would not interfere with the quality of service they come for. Laboratory results were communicated to the physician according to turnaround time for proper patient management.

### Study population

The study population was patients of all age groups presenting with middle ear infections attended at KCMC, ENT department. The study included all patients of all ages attended at KCMC, ENT department presenting with middle ear infections at the prescribed study period. The study excludes Patients referred to KCMC, ENT Department with middle ear infections but on to topical medications, those with post-ear surgery CSOM-related discharge i.e., tympan mastoidectomy or canaloplasty, and patients of all ages attended at KCMC, ENT department presenting with middle ear infections but refuse to consent to participate in the study.

### Sample size and sampling technique

Daniel’s formula was used to estimate the minimum sample size given the unknown standard deviation and the proportion of CSOM at KCMC.

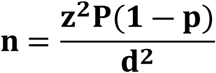

Where, n = minimum sample size, Z = 1.96 at 95% Confidence interval, P = Prevalence of CSOM in developing countries, P =14 % (32), d= acceptable marginal error =0.05. 10% of the calculated sample size was added to cover any rejected samples. The calculated minimum sample size was 201 participants. A convenient sampling method was used to obtain study participants who consented to participate in the study until the desired minimum sample size was reached. The method was conducted using a prescribed consent form, (S1 Appendix).

### Data collection tools, methods, procedures, and identification

Participants’ demographic and clinical information was collected by using standardized data collection tool, (S2 Appendix) while laboratory information was collected by using laboratory worksheets, (S3 Appendix).

During sample collection, an ENT specialist examined the participants’ middle ear/tympanic membrane with a powered otoscope or ear microscope for perforation. If clinical features were confirmed to have CSOM, a sterile swab was used to collect a middle ear discharge sample. With the aid of an ear speculum, the external ear canal was bypassed to avoid contamination, and one discharge swab was collected from each affected middle ear cavity, placed into Copan eswab transport media, and transported to the Laboratory for culture and sensitivity within less than two hours after collection (7,33).

All biological samples were sent to the laboratory and processed in biological safety cabinet level II. The gram staining was conducted for each sample and observed microscopically for the presence of bacteria and their gram reactions (34). Each specimen was cultured into Blood Agar (BA), MacConkey Agar (MCA), Chocolate agar (CA), and Sabouraud Dextrose Agar (SDA) and incubated aerobically at 37 degrees Celsius for 18-24 hours except for CA which was incubated while placed in the candle jar at mentioned temperature and time. The plates with no growth were re-incubated for up to 48 hours and if there was still no growth, it was reported as ‘no bacterial growth’ and discarded. All plates with Bacterial growth were identified by standard microbiological methods depending on culture observable features and gram reaction (35,36).

### Bacteria identification

Bacteria were identified using various conventional microbiological identification tests such as gram staining reaction, culture characteristics, enzyme production, utilization of various substrates, and production of metabolites (37). For gram-negative bacteria oxidase TSI, SIM, citrate, indole, and urease biochemical tests were used. Gram-positive bacteria were identified based on catalase and coagulase biochemical tests as shown (Figures 1, 2, and 3) (38). Laboratory information was collected by using a laboratory work sheet, (S3 Appendix).

**Figure 1:**
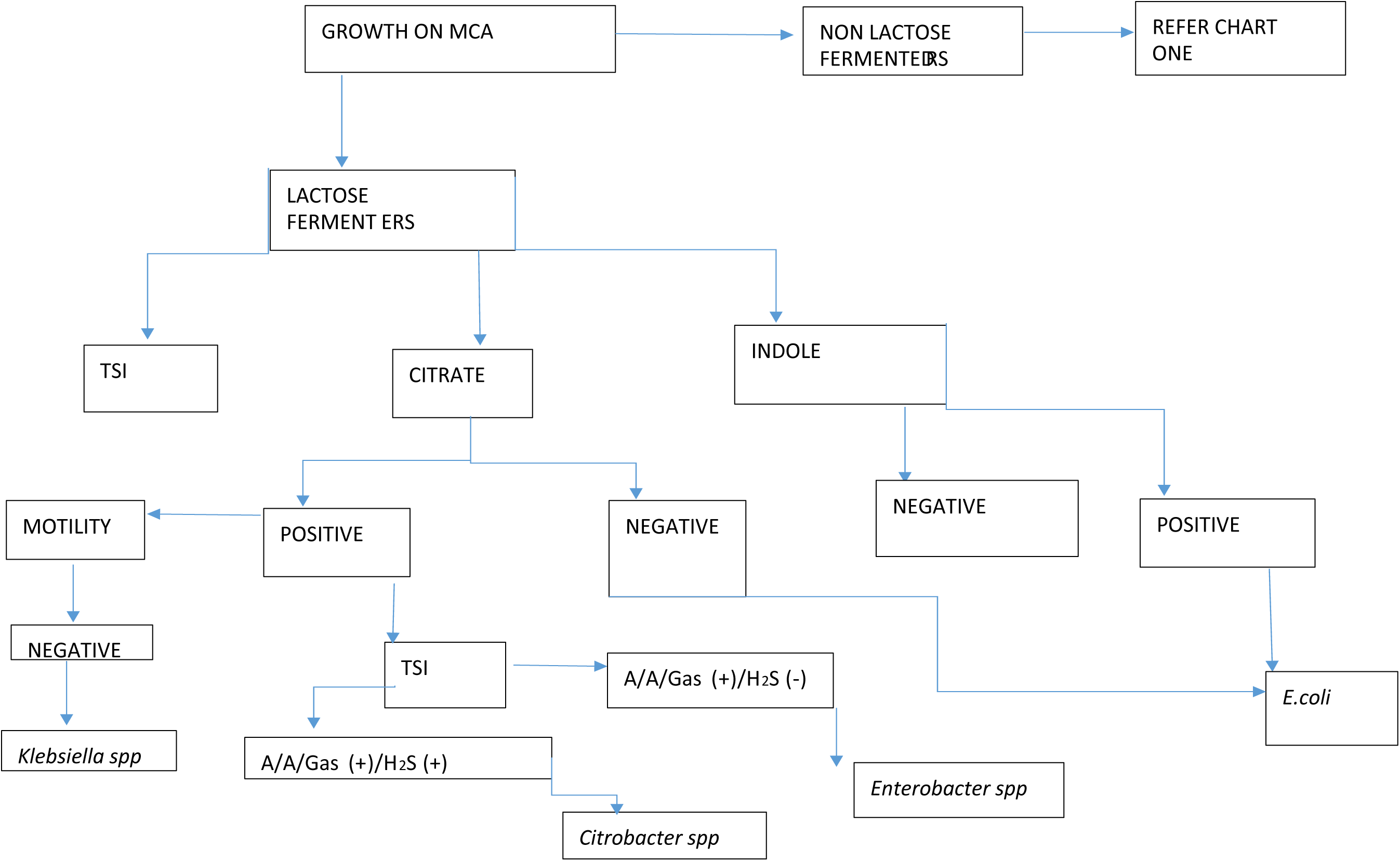
Identification chart flow for gram-negative rods.

**Figure 2:**
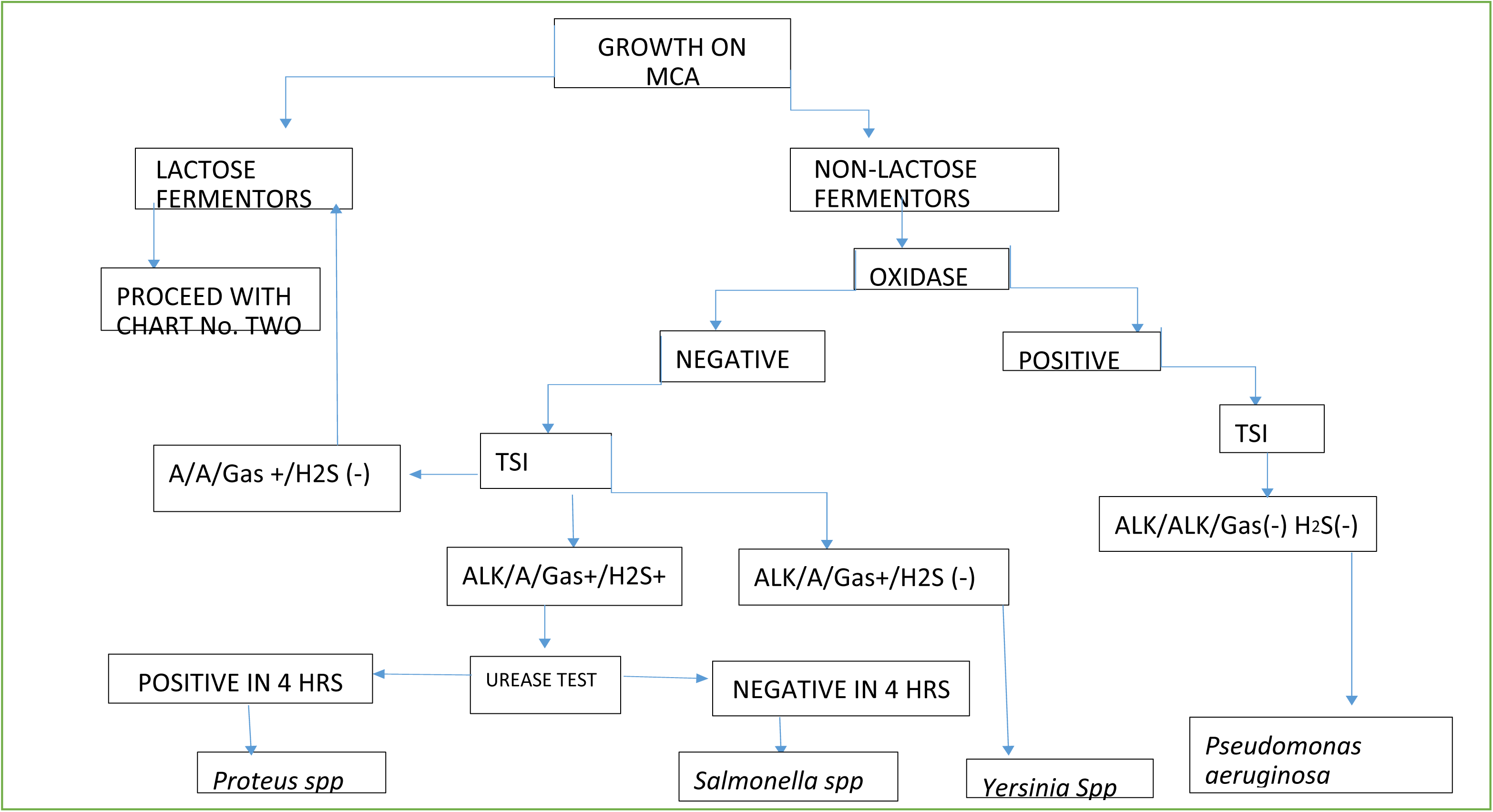
Identification chart flow for gram-negative rods.

**Figure 3:**
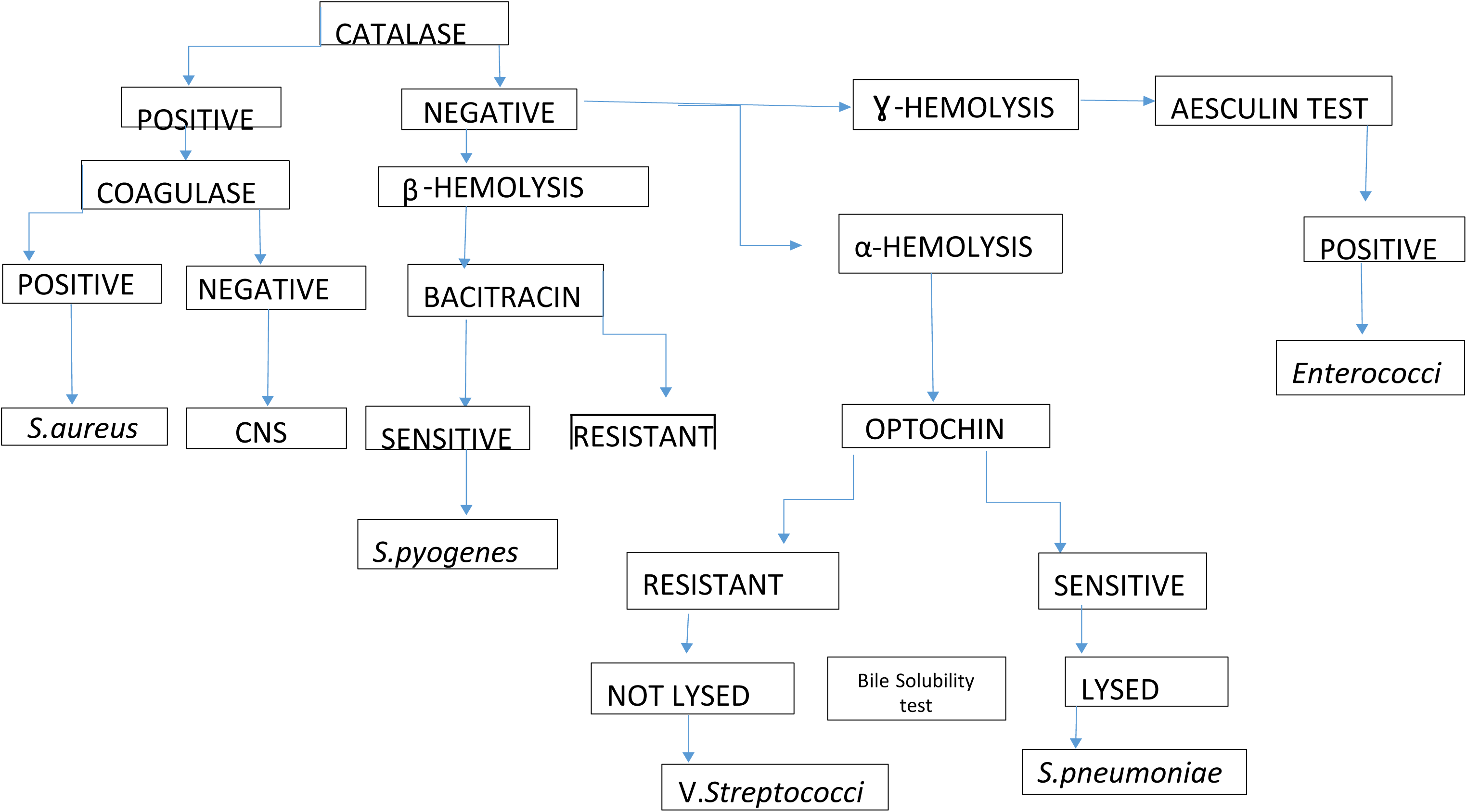
Identification chart flow for gram-positive cocci.

### Drug Susceptibility testing

Kirby Bar disc diffusion method was used to test the antimicrobial susceptibility pattern of the purely isolated and identified bacteria according to CLSI standard. A small portion of Bacterial colonies was suspended in physiological saline to make a concentration equivalent to 0.5 McFarland’s standards and pre-incubated for 15 minutes at 35 degrees Celsius to attain fluent growth. The suspension was then rubbed on the surface of Mueller and Hinton agar and incubated at 35 degrees Celsius for 18 hours. The zones of inhibition were measured to the nearest millimeters using a vernier caliper. The results were reported as sensitive or resistant on the drug susceptibility test worksheet, (S4 Appendix), according to CLSI guidelines (39).

### Statistical analysis

Obtained data were checked manually for completeness and correctness on the Data collection tools sheet on daily basis before entering into computer software. SPSS software version 21 (IBM Corp, Chicago IL) was used to clean and analyze the data. Frequencies and proportions were used to summarize data and presented using tables, graphs, Charts and figures.

## RESULTS

### Prevalence of Chronic Suppurative Otitis

Overall, of all 201 participants enrolled in this study, the prevalence of chronic supportive Otitis was 11.4% (23/201) as shown in Figure 4 below.

**Figure 4:**
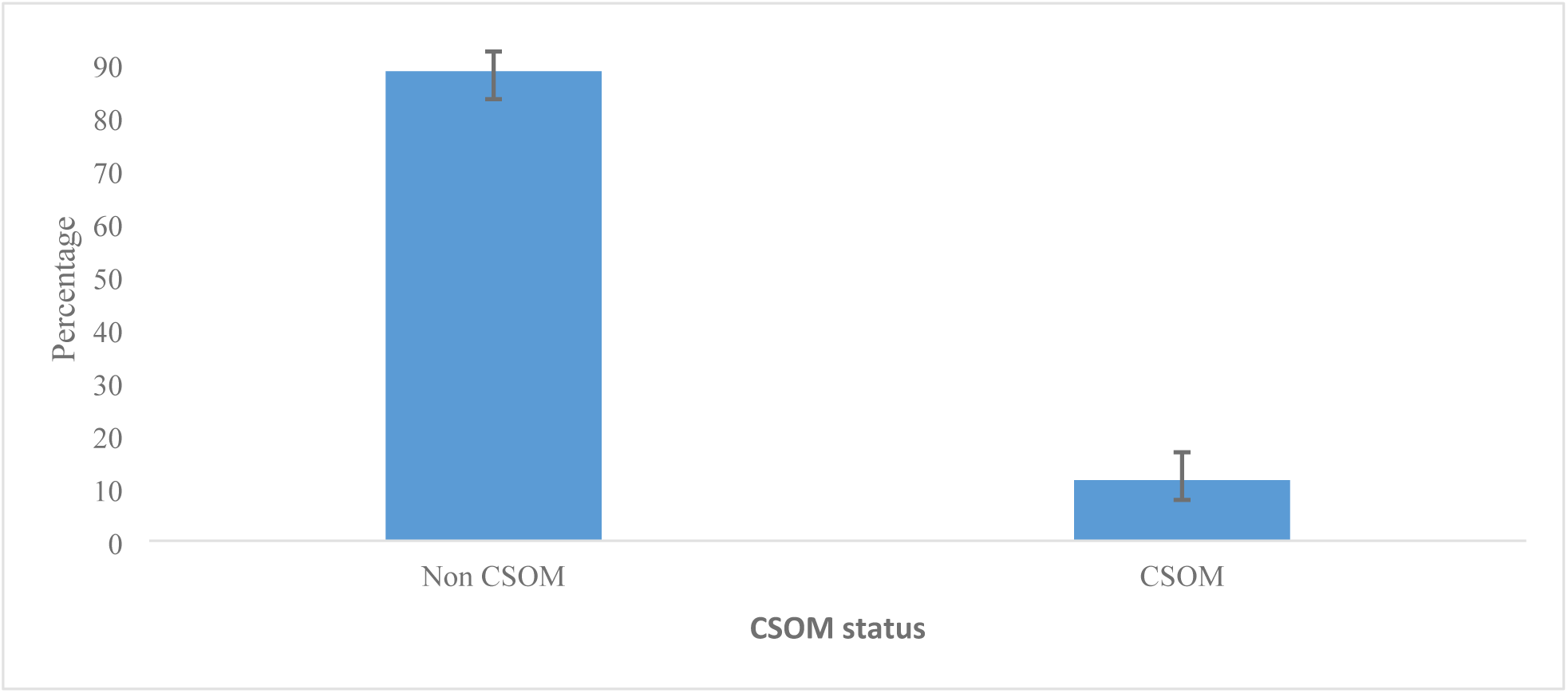
Prevalence of CSOM.

### Sociodemographic and clinical characteristics of study participants

In total, 201 participants were enrolled in the study during (March to May 2022). The majority of participants 57.7% (116/201) were female. A high proportion of the study participants were above 15 years of age 56.7% (114/201), and the median age was 18 years with an Interquartile range (5–28). The majority 53.7% (108/201) of the total participants were residing in rural areas. Of all participants, only 19.4% (39/201) had upper respiratory tract infections, table 1.

**Table 1:** Sociodemographic and clinical characteristics of study participants by CSOM (N=201)

| Characteristics | Total | Chronic Suppurative Otitis |  | p-value |
| --- | --- | --- | --- | --- |
|  | n (%) | n (%) |  |  |
|  |  | No | Yes |  |
| Age group (in years) |  |  |  | 0.003 |
| <5 | 58 (28.9) | 45 (77.6) | 13 (22.4) |  |
| 5-15 | 29 (14.4) | 25 (86.2) | 4 (13.8) |  |
| 16+ | 114(56.7) | 108 (94.7) | 6 (5.3) |  |
| Median (Inter quartile range) | 18 (5-28) |  |  |  |
| Sex |  |  |  | 0.568 |
| Male | 85 (57.70 | 74 (87.1) | 11 (12.9) |  |
| Female | 116 (57.71) | 104 (89.7) | 12 (10.3) |  |
| Residence |  |  |  | 0.039 |
| Rural | 108 (53.7) | 91 (84.3) | 17 (15.7) |  |
| Urban | 93 (46.3) | 87 (93.6) | 6 (6.4) |  |
| Upper respiratory infections |  |  |  | 0.166 |
| No | 162 (80.6) | 146 (90.1) | 16 (9.9) |  |
| Yes | 39 (19.4) | 32 (82.1) | 7 (17.9) |  |

During the chi-square test, age and residence were significantly associated with chronic suppurative Otitis infections. The prevalence of CSOM in age groups was 22.4% (13/58) under 5 years, 13.8% (4/29) in 5-15 years, and 5.3% (6/114) in above 15 years. Regarding sex, the proportion of CSOM infection was higher among males 12.9% (11/85) compared to females 10.3% (12/116). In addition, there was a high proportion of CSOM cases 15.7% (17/108) among rural participants, table 1.

### Microbiological profile of bacteria isolated from CSOM patients

For all participants diagnosed with CSOM infections (n=23), a Microbiological culture was performed to identify the bacteriological profile and Culture positive was 82.6% (n=19). *S aureus* had the highest proportion of 28.57%, followed by *Pseudomonas aeruginosa* 19.04%, *Proteus spp* 14.14%, *Coagulase Negative Staphylococci, E coli,* and *Enterobacter spp 9.52*% each, *H influenzae* and *Citrobacter sp* 4.16% each, figure 5.

**Figure 5:**
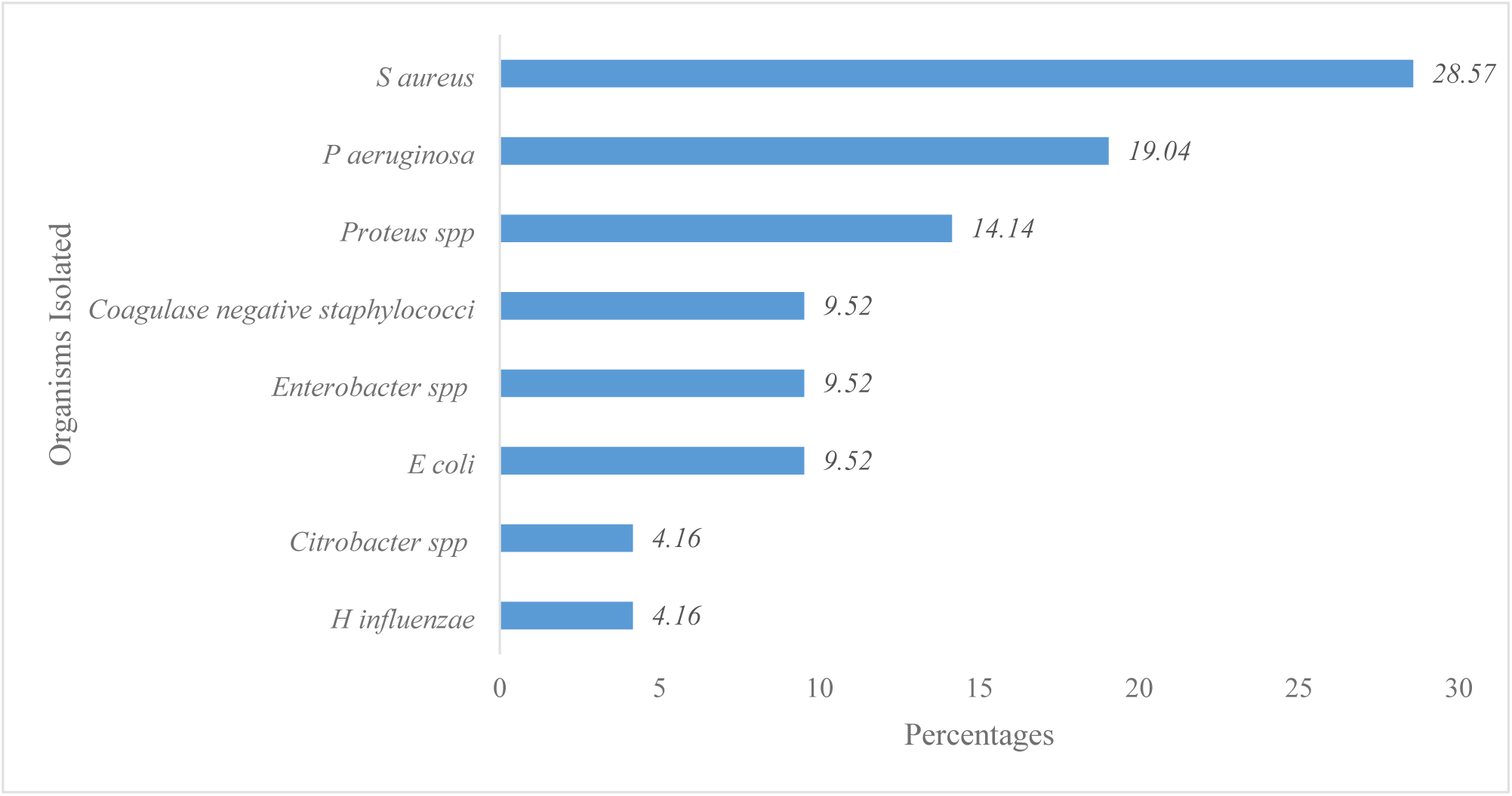
Proportion of microbiological culture among CSOM-positive participants.

### Antimicrobial susceptibility pattern of CSOM bacterial isolates

Twenty different bacterial isolates from CSOM discharge were tested against 14 antibiotics according to CLSI guidelines. Highest drug resistance was observed against Amoxicillin/Clavulanate 70%, Ampicillin 60%, Cefoxitin 50% while lower resistance proportion were observed in Cefotaxime 11.11%, Meropenem 12.5%, Imipenem 18.18% Ciprofloxacin 15.79%, Chloramphenicol 18.75%, Ceftazidime 33.3% and Erythromycin 37.5%. There was no resistance to Amikacin, figure 6.

**Figure 6:**
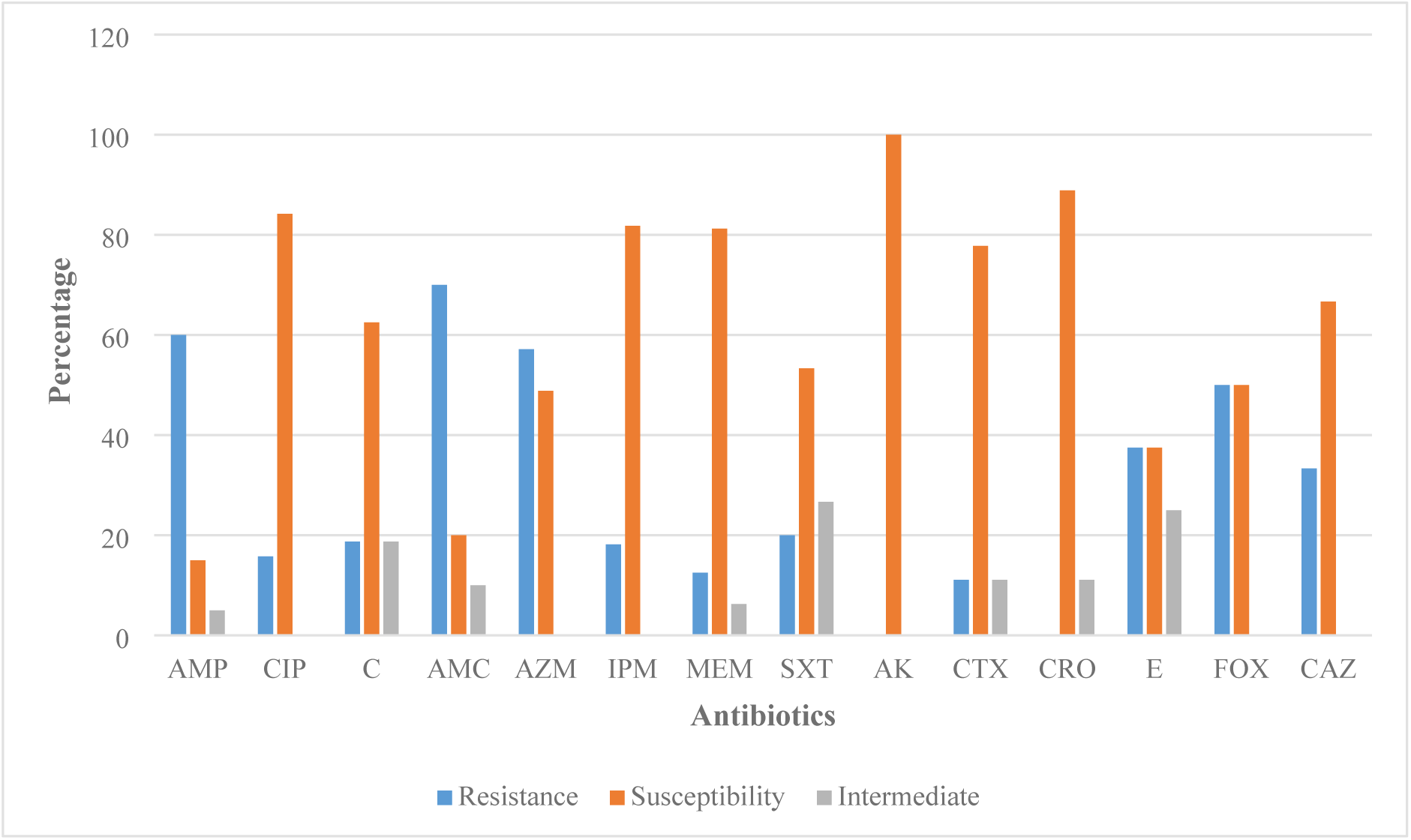
Antimicrobial susceptibility pattern among CSOM bacterial isolates.

## DISCUSSION

### Prevalence of Chronic Suppurative Otitis Media

The prevalence of CSOM varies across various geographical locations, ethnic groups, and various social practices (7,13). This study found an 11.4% prevalence of CSOM at KCMC hospital, which is categorized as a high CSOM prevalence by WHO (7). Another study conducted in Dar es Salaam reported similar results (40). In 2004 WHO reported Tanzania among the countries with the highest CSOM prevalence (14%), which is slightly higher than our findings. Other studies conducted in Tanzania had lower prevalences 2.6% and 1.4% (41,42) respectively, and the lower prevalence in these studies might have been due to the majority of patients attending Muhimbili where the studies were conducted coming from urban areas while most of the people served by KCMC where this study was conducted are from rural areas, as it has been shown by many studies, including our study there is high CSOM prevalence among rural residents compared to urban. Another study conducted in northern Tanzania reported a lower prevalence of 1.6% (43), but this difference could be because it was a community-based study while our study is hospital-based. Age and living in rural areas were significantly associated with chronic suppurative Otitis infections (p-value=0.003, 0.039 respectively). 22.4% of CSOM cases were participants aged below five years which is similar to other studies (44,45). Our study shows that 15.7% of rural participants had CSOM and only 6.4% among urban participants. Another study done in Dar es Salaam also report**ed** higher CSOM prevalence among rural participants compared to urban participants (32). Gender and Upper respiratory tract infections were associated with chronic suppurative Otitis infections but not statistically significant (p-value=0.58, 0.166 respectively). The proportion of CSOM infection was higher among males (12.9%) compared to females (10.3%). Another study conducted in Dar es Salaam reported a predominance of CSOM in males compared to females (46). Nearly similar results were also reported in India (5).

### Microbiological profile of CSOM Isolates

The predominance of Bacterial species in CSOM patients is predetermined by various factors such as geographical location, antibiotic usage, lifestyle such as overcrowding, poor hygiene, and swimming in pods among others. Bacteria can enter the middle ear from various sources like the environment, infection from other persons, and indigenously from URT through the Eustachian tube.

In the current study, *S aureus* was the most common isolate with a proportion of 28.57%, followed by *Pseudomonas aeruginosa* 19.04%, *Proteus spp* 14.14%, *Coagulase Negative Staphylococci, E coli,* and *Enterobacter spp 9.52*% each, *H influenzae* and *Citrobacter sp* 4.16% each. A study conducted in Dar es Salaam Tanzania gives a similar picture (47). The predominance of *S aureus* and *Pseudomonas aeruginosa* in CSOM patients is in line with other studies conducted in Ethiopia, India, and Australia (3,18,20,48,49). Bacterial isolates were predominantly gram-negative (60%) similar to other studies conducted in Dar es Salaam (46), Ethiopia (18,50), and India (19,51). *Pseudomonas aeruginosa* was the most common isolate among gram-negative rods in our study, though the coliform group when combined had the highest proportion among all isolates. While this was similar to other studies in Africa (18,50), a study conducted in Dar es Salaam showed different results with *Klebsiella pneumonia, Proteus spp*, and *E coli* as the predominant isolates from CSOM infections (46). The high proportion of coliform isolates from CSOM patients in our study could be due to unhygienic practices such as swimming in contaminated ponds and improper hand washing among patients who attended KCMC. Among gram-positive *S aureus* had a higher proportion than Coagulase Negative *Staphylococci,* similar to results reported by Hailu (16). In contrast to our study, Kaur reported Streptococci as the second common isolate (52), and this difference in findings can be due to differences in geographical location, lifestyle of study participants, and prevalence of Upper respiratory tract infections. In this study, the poly-microbial infection was 9.52%, though most studies, including Abraham et al in Dar es Salaam, show higher proportions of poly-microbial in CSOM patients (18,42). The use of wide-spectrum quinolones at KCMC might have reduced bacterial CSOM co-infections. Biofilms shield bacteria from antimicrobials (53), and due to its ability to form biofilm, *Pseudomonas aeruginosa* poses a threat of treatment failure and possible complications to many CSOM patients considering *Pseudomonas aeruginosa* being the second common isolate in our study. A biofilm-forming pathogen may be reported as susceptible to a particular antibiotic in laboratory analysis but the biofilm formation in the middle ear may prevent the effective concentration of antibiotic from reaching some bacterial cells resulting in treatment failure. This could also be the reason why this bacterium is common in CSOM infections despite the wide spectrum of topical antimicrobial use.

### Antibiotic susceptibility pattern of bacteria isolated from CSOM patients

CSOM is normally treated with a cocktail of topical antifungals, antibiotics, and antiseptics. In this study, we concentrated on Bacteria and their susceptibility to common antimicrobials used at KCMC hospital.

Twenty different bacterial isolates from CSOM discharge were tested against 14 antibiotics according to CLSI guidelines. The highest drug resistance was observed against the penicillin class of antibiotics Amoxicillin/Clavulanate 70% and Ampicillin 60%. This agrees with other studies with similar bacterial agents. A study conducted in Dar es Salaam presents similar findings (46). In Ethiopia, it was reported resistance of 88.3% and 79.5% respectively. Also, a study conducted in Sab Sahara Africa presents similar results (54). Gram-positive cocci had a resistance of 50% to Cefoxitin, this signifies a high predominance of Methicillin-Resistant *Staphylococci* (MRSA) in CSOM infections. Several other studies report the presence of MRSA CSOM (5,51). This study found a lower resistance proportion in Cefotaxime 11%, Meropenem 12.5%, Ciprofloxacin 15.79%, Chloramphenicol 18.75%, Ceftazidime 33.3% and Erythromycin 37.5% while there was no resistance to Amikacin in this study. This is in line with other studies in Dar es Salaam, India, and Sab Sahara Africa with etiological agents of the same nature (46,54,55). Resistance of 15.79% against Ciprofloxacin should not be overlooked as it is the most desirable topical antibiotic due to its effectiveness and low ototoxicity, it is also a recommended topical antibiotic by WHO and Tanzania treatment guideline manual (7,28). All bacterial isolates were resistant to more than one antibiotic, this gives a clear picture of Multidrug resistance (MDR) among CSOM bacteria. Similar to these many studies isolated MDR bacteria in CSOM patients (23,27). Macrolides and aminoglycosides had lower resistance levels but their use has clinical implications for vestibulotoxicity and Cochleotoxicity (6).

## Conclusion

The study reported a high prevalence of CSOM and a high proportion of MDR bacteria among CSOM patients who attended KCMC hospitals. Continuing empirical treatment could escalate the problem and complicated sequelae. Further studies at zonal and country levels are recommended to establish an updated burden of CSOM in Tanzania. We also highly recommend microbiological analysis of middle ear discharge to study bacterial profile and drug susceptibility to provide effective antibiotics for treatment. An antibiotic stewardship program for surveillance of antimicrobial resistance in CSOM bacterial isolates should be developed at KCMC Hospital.

## Acknowledgments

The authors would like to thank the management of KCMC Hospital for permission to conduct the study as well as the KCMC University for their permission to use its facility to conduct the study.

## Authors’ contributions

**SM** contributed to Conceptualization, Data Curation, Formal Analysis, Investigation, Methodology, Resources, Software, Validation, Visualization, Writing – Original Draft Preparation, Writing – Review & Editing.

**DM**: contributed to Conceptualization, Data Curation, Formal Analysis, Investigation, Methodology, Resources, Software, Validation, Visualization, Writing – Original Draft Preparation, Writing – Review & Editing.

**PT**: contributed to Data Curation Project Administration, Formal Analysis, Investigation, Methodology, Resources, Software, Validation, Visualization, Writing – Original Draft Preparation, Writing – Review & Editing.

**HJM, JN & DJO:** Formal Analysis, Investigation, Methodology, Resources, Software, Validation, Visualization, Writing – Original Draft Preparation, Writing – Review & Editing

**EBG & AM:** Contributed in Conceptualization, Data Curation, Formal Analysis, Funding Acquisition, Investigation, Methodology, Project Administration, Resources, Validation, Visualization, Writing – Original Draft Preparation, Writing – Review & Editing

**DCK**: Contributed in Supervision, Data Curation, Formal Analysis, Funding Acquisition, Investigation, Methodology, Project Administration, Resources, Validation, Visualization, Writing – Original Draft Preparation, Writing – Review & Editing

All authors read and approved the final manuscript.

## Data availability

The datasets used and/or analyzed during the current study are available from the corresponding author upon reasonable request.

## Consent for publication

NA

## REFERENCES

1. Adoga AS, Màan EN, Malu D, Badung BP, Obiesie I V., Nwaorgu OGB. Swab and aspiration specimen collection methods and antibiogram in chronic suppurative otitis media at Jos University Teaching Hospital: Which is superior. Ann Afr Med. 2010;9(4):230–4.

2. Bluestone CD. Epidemiology and pathogenesis of chronic suppurative otitis media : implications for prevention and treatment 1. 1998;42(November 1996):207–23.

3. Rosario DC MMD. Chronic Suppurative Otitis. Treasure Island (FL): StatPearls Publishing [Internet]. 2021 [cited 2021 Dec 29]; Available from: https://www.ncbi.nlm.nih.gov/books/NBK554592/

4. WHO. Chronic suppurative otitis media : burden of illness and management options [Internet]. Geneva PP - Geneva: World Health Organization; 2004. Available from: https://apps.who.int/iris/handle/10665/42941

5. Kumar Shetty A, Majumder P, Bhat Tulasidas M. Microbial profile of ear discharge in children with chronic suppurative otitis media and the antibiotic susceptibility pattern of bacterial isolates. Indian Journal of Microbiology Research. 2019 Jun 28;6(2):174–9.

6. Mittal R, Lisi C V, Gerring R, Mittal J, Mathee K, Narasimhan G, et al. Current concepts in the pathogenesis and treatment of chronic suppurative otitis media. 2015;1103–16.

7. WHO. Chronic suppurative otitis media : burden of illness and management options. Geneva PP - Geneva: World Health Organization; 2004.

8. Mendez R and. Chronic Suppurative Otitis. Treasure Island (FL): StatPearls Publishing. 2021;

9. Rovers MM. The burden of otitis media. Vaccine. 2008;26(SUPPL. 7):2–8.

10. WHO. 1 in 4 people is projected to have hearing problems by 2050. Geneva; 2021.

11. Libwea JN, Kobela M, Ndombo PK, Syrjänen RK, Huhtala H, Fointama N, et al. The prevalence of otitis media in 2–3-year-old Cameroonian children estimated by tympanometry. Int J Pediatr Otorhinolaryngol. 2018;115(June):181–7.

12. Riaz N, Rehman AU, Sheikh AH, Aslam N, Malik I, Riaz B. Frequency of sensorineural hearing loss in chronic suppurative otitis media. Pakistan Journal of Medical and Health Sciences. 2018;12(4):1635–7.

13. Monasta L, Ronfani L, Marchetti F, Montico M, Brumatti L, Bavcar A, et al. Burden of disease caused by otitis media: Systematic review and global estimates. PLoS One. 2012;7(4).

14. Choffor-Nchinda E, Bola Siafa A, Nansseu JR. Otitis media with effusion in Africa-prevalence and associated factors: A systematic review and meta-analysis. Laryngoscope Investig Otolaryngol. 2020;5(6):1205–16.

15. Li MG, Hotez PJ, Vrabec JT, Donovan DT. Is Chronic Suppurative Otitis Media a Neglected Tropical Disease? PLoS Negl Trop Dis. 2015;9(3):1–6.

16. Hailu D, Mekonnen D, Derbie A, Mulu W, Abera B. Pathogenic bacteria profile and antimicrobial susceptibility patterns of ear infection at Bahir Dar Regional Health Research Laboratory Center, Ethiopia. Springerplus. 2016;5(1).

17. Adoga AS, Màan EN, Malu D, Badung BP, Obiesie I V., Nwaorgu OGB. Swab and aspiration specimen collection methods and antibiogram in chronic suppurative otitis media at Jos University Teaching Hospital: Which is superior. Ann Afr Med. 2010;9(4):230–4.

18. Wasihun AG, Zemene Y. Bacterial profile and antimicrobial susceptibility patterns of otitis media in Ayder Teaching and Referral Hospital, Mekelle University, Northern Ethiopia. Springerplus. 2015 Dec 1;4(1):1–9.

19. Hiremath B, Mudhol RS, Vagrali MA. Bacteriological Profile and Antimicrobial Susceptibility Pattern in Chronic Suppurative Otitis Media: A 1-Year Cross-Sectional Study. Indian Journal of Otolaryngology and Head and Neck Surgery [Internet]. 2019 Nov 1;71(s2):1221–6. Available from: 10.1007/s12070-018-1279-6

20. Khatun MR, Alam KMF, Naznin M, Salam MA. Microbiology of chronic suppurative Otitis media: An update from a tertiary care hospital in Bangladesh. Pak J Med Sci. 2021;37(3):821–6.

21. S H C, M M K, Handi P, Khavasi P, Doddmani SS, Riyas M. To Study the Level of Awareness About Complications of Chronic Suppurative Otitis Media (CSOM) in CSOM Patients. J Clin Diagn Res. 2014 Feb;8(2):59–61.

22. Park DC, Lee SK, Cha CI, Lee S. Antimicrobial resistance of Staphylococcus from otorrhea in chronic suppurative otitis media and comparison with results of all isolated Staphylococci. 2008;571–7.

23. Chong LY, Head K, Richmond P, Snelling T, Schilder AGM, Burton MJ, et al. Systemic antibiotics for chronic suppurative otitis media. Cochrane Database of Systematic Reviews. 2018 Jun;2018(6):CD013052.

24. WHO. Antimicrobial resistance. 2021.

25. Mboya EA, Davies ML, Horumpende PG, Ngocho JS. Inadequate knowledge of appropriate antibiotic use among clients in the Moshi municipality Northern Tanzania. PLoS One. 2020;15(9 September):1–13.

26. Mund MD, Khan UH, Tahir U, Mustafa BE, Fayyaz A. Antimicrobial drug residues in poultry products and implications on public health: A review. Int J Food Prop. 2017;20(7):1433–46.

27. Lee SK, Park DC, Kim MG, Boo SH, Choi YJ, Byun JY, et al. Rate of isolation and trends of antimicrobial resistance of multidrug-resistant pseudomonas aeruginosa from otorrhea in chronic suppurative otitis media. Clin Exp Otorhinolaryngol. 2012;5(1):17–22.

28. NMTC. Standard Treatment Guidelines and Essential Medicines List for. Sixth edit. Vol. Sixth edit. Dodoma: The United Republic of Tanzania standard treatment guidelines and national essential medicines list for Tanzania’s mainland ministry of health, community development, gender, elderly, and children.; 2021.

29. Gorems K, Beyene G, Berhane M, Mekonnen Z. Antimicrobial susceptibility patterns of bacteria isolated from patients with ear discharge in Jimma Town, Southwest, Ethiopia. BMC Ear Nose Throat Disord. 2018;18(1):1–9.

30. Khatun MR, Alam KMF, Naznin M, Salam MA. Microbiology of chronic suppurative Otitis media: An update from a tertiary care hospital in Bangladesh. Pak J Med Sci. 2021;37(3):821–6.

31. Bell BG, Schellevis F, Stobberingh E, Goossens H, Pringle M. A systematic review and meta-analysis of the effects of antibiotic consumption on antibiotic resistance. BMC Infect Dis. 2014;14(1):1–25.

32. Minja BM, Machemba A. Prevalence of otitis media, hearing impairment and cerumen impaction among school children in rural and urban Dar es Salaam, Tanzania. Int J Pediatr Otorhinolaryngol. 1996;37(1):29–34.

33. Public Health England. UK Standards for Microbiology Investigation of Ear Infections and Associated Specimens Investigation of Ear Infections and Associated Specimens. 2014;1–18.

34. Smith ACMAH. Gram Staining Protocols. American Society For Microbiology. 2005;

35. Leber, American Society for Microbiology, AL. Clinical microbiology procedures handbook. 2016.

36. Public Health England. UK Standards for Microbiology Investigation of Ear Infections and Associated Specimens Investigation of Ear Infections and Associated Specimens. 2014;1–18.

37. Srivastava VK. Standard Operating Procedures Bacteriology, Antimicrobial Resistance Surveillance and Research Network. 2015;1–160.

38. Cheesbrough M. District Laboratory Practice in Tropical Countries. 2nd ed. Cambridge: Cambridge University Press; 2006.

39. Cappuccino JG, Sherman N. Microbiology : a laboratory manual. 9th ed. San Francisco: Benjamin-Cummings; 2011.

40. Manni JJ, Lema PN. Otitis media in Dar es Salaam, Tanzania. J Laryngol Otol [Internet]. 2007/06/29. 1987;101(3):222–8. Available from: https://www.cambridge.org/core/article/otitis-media-in-dar-es-salaam-tanzania/B0AAE0ADD1FED31919E1F79408BC8D6C

41. Minja BM, Machemba A. Prevalence of otitis media, hearing impairment and cerumen impaction among school children in rural and urban Dar es Salaam, Tanzania. Int J Pediatr Otorhinolaryngol. 1996;37(1):29–34.

42. Abraham ZS, Ntunaguzi D, Kahinga AA, Mapondella KB, Massawe ER, Nkuwi EJ, et al. Prevalence and etiological agents for chronic suppurative otitis media in a tertiary hospital in Tanzania. BMC Res Notes [Internet]. 2019;12(1). Available from: 10.1186/s13104-019-4483-x

43. Bastos I, Mallya J, Ingvarsson L, Reimer Å, Andréasson L. Middle ear disease and hearing impairment in northern Tanzania. A prevalence study of schoolchildren in the Moshi and Monduli districts. Int J Pediatr Otorhinolaryngol. 1995;32(1):1–12.

44. Monasta L, Ronfani L, Marchetti F, Montico M, Brumatti L, Bavcar A, et al. Burden of disease caused by otitis media: Systematic review and global estimates. PLoS One. 2012;7(4).

45. Choffor-Nchinda E, Bola Siafa A, Nansseu JR. Otitis media with effusion in Africa-prevalence and associated factors: A systematic review and meta-analysis. Laryngoscope Investig Otolaryngol. 2020;5(6):1205–16.

46. Abraham ZS, Ntunaguzi D, Kahinga AA, Mapondella KB, Massawe ER, Nkuwi EJ, et al. Prevalence and etiological agents for chronic suppurative otitis media in a tertiary hospital in Tanzania. BMC Res Notes. 2019;12(1).

47. Moshi NH, Minja BM, Ole-Lengine L, Mwakagile DS. Bacteriology of chronic otitis media in Dar es Salaam, Tanzania. East Afr Med J. 2000 Jan;77(1):20–2.

48. Couzos S, Lea T, Mueller R, Murray R, Culbong M. Effectiveness of to topical antibiotics for chronic suppurative otitis media in Aboriginal children: A community-based, multicentre, double-blind randomized controlled trial. Medical Journal of Australia. 2003;179(4):185–90.

49. Lee SK, Park DC, Kim MG, Boo SH, Choi YJ, Byun JY, et al. Rate of isolation and trends of antimicrobial resistance of multidrug resistant pseudomonas aeruginosa from otorrhea in chronic suppurative otitis media. Clin Exp Otorhinolaryngol. 2012;5(1):17–22.

50. Hailu D, Mekonnen D, Derbie A, Mulu W, Abera B. Pathogenic bacteria profile and antimicrobial susceptibility patterns of ear infection at Bahir Dar Regional Health Research Laboratory Center, Ethiopia. Springerplus. 2016 Dec 1;5(1).

51. Kaur DrP, Singh Sood DrA, Sharma DrS, Awal DrG. Microbiological profile and antimicrobial susceptibility pattern of chronic suppurative otitis media in a tertiary care centre. Tropical Journal of Pathology and Microbiology. 2018;4(1):1–13.

52. Kaur DrP, Singh Sood DrA, Sharma DrS, Awal DrG. Microbiological profile and antimicrobial susceptibility pattern of chronic suppurative otitis media in a tertiary care centre. Tropical Journal of Pathology and Microbiology. 2018;4(1):1–13.

53. Dohar JE, Hebda PA, Veeh R, Awad M, Costerton JW, Hayes J, et al. Mucosal Biofilm Formation on Middle-Ear Mucosa in a Nonhuman Primate Model of Chronic Suppurative Otitis Media. 2005;(August):1469–72.

54. Tesfa T, Mitiku H, Sisay M, Weldegebreal F, Ataro Z, Motbaynor B, et al. Bacterial otitis media in sub-Saharan Africa: A systematic review and meta-analysis. BMC Infect Dis. 2020;20(1):1–12.

55. Shetty AK, Majumder P, Bhat Tulasidas M. Microbial profile of ear discharge in children with chronic suppurative otitis media and the antibiotic susceptibility pattern of bacterial isolates. Indian Journal of Microbiology Research. 2019;6(2):174–9.

